# Toward a COVID-19 testing policy: where and how to test when the purpose is to isolate silent spreaders

**DOI:** 10.1101/2020.12.22.20223651

**Authors:** AL Rivas, AL Hoogesteijn, JB Hittner, MHV van Regenmortel, P Kempaiah, P Vogazianos, A Antoniades, A Ioannidis, JL Febles, FO Fasina

## Abstract

**Background:** To stop pandemics, such as COVID-19, infected individuals should be detected, treated if needed, and –to prevent contacts with susceptible individuals-isolated. Because most infected individuals may be asymptomatic, when testing misses such cases, epidemics may growth exponentially, inducing a high number of deaths. In contrast, a relatively low number of COVID-19 related deaths may occur when both symptomatic and asymptomatic cases are tested.

**Methods:** To evaluate these hypotheses, a method composed of three elements was evaluated, which included: (i) county- and country-level geo-referenced data, (ii) cost-benefit related considerations, and (iii) temporal data on mortality or test positivity (TP). TP is the percentage of infections found among tested individuals. Temporal TP data were compared to the tests/case ratio (T/C ratio) as well as the number of tests performed/million inhabitants (tests/mi) and COVID-19 related deaths/million inhabitants (deaths/mi).

**Findings:** Two temporal TP profiles were distinguished, which, early, displayed low (∼ 1 %) and/or decreasing TP percentages or the opposite pattern, respectively. Countries that exhibited >10 TP % expressed at least ten times more COVID-19 related deaths/mi than low TP countries. An intermediate pattern was identified when the T/C ratio was explored. Geo-referenced, TP-based analysis discovered municipalities where selective testing would be more cost-effective than alternatives.

**Interpretations:** When TP is low and/or the T/C ratio is high, testing detects asymptomatic cases and the number of COVID-19 related deaths/mi is low. Geo-referenced TP data can support cost-effective, site-specific policies. TP promotes the prompt cessation of epidemics and fosters science-based testing policies.

**Funding:** None

**Research in context:** *Evidence before this study:* To map this field, bibliographic searches were conducted in the *Web of Science*, which included the following results: (i) COVID-19 (95,133 hits), (ii) SARS COV-2 (33,680 hits), (iii) testing policy and COVID-19 (939 hits), (iv) testing policy and SARS COV-2 (340 hits), (v) testing policy and COVID-19 and asymptomatic (80 hits), (vi) testing policy and SARS COV-2 and asymptomatic (54 hits); (vii) test positivity and COVID-19 and validation (7 hits), and (viii) test positivity and SARS CoV-2 and validation (5 hits). Therefore, before this study, testing policy in relation to asymptomatic cases as well as test positivity represented a very low proportion (between ∼1 thousandth to ∼ 1 ten thousandth) of all publications. While many articles distinguished between diagnostic and screening tests, no paper was found in which testing policy is mentioned as part of a process ultimately designed to isolate all infected individuals. The few articles that mentioned test positivity only investigated symptomatic cases. These quanti/qualitative assessments led the authors to infer that neither testing policy nor test positivity had been adequately validated and/or investigated.

*Added value of this study:* We provide the first validation of test positivity as an estimate of disease prevalence under rapidly changing conditions: in pandemics, disease prevalence may vary markedly within short periods of time. We also address a double limitation of control campaigns against COVID-19, namely: it is unknown *who* and *where* to test. Asymptomatic cases are not likely to seek medical assistance: while they feel well, they silently spread this pandemic. Because they represent approximately half of all infected individuals, they are a large, moving, and invisible target. *Where* to find them is also unknown because (i) randomized testing is likely to fail and (ii) testing is very limited. Usually, the locations where infected people reside are not randomly distributed but geographically clustered, and, up to now less than four persons per thousand inhabitants are tested on a given day. However, by combining geo-referenced test positivity data with cost-benefit considerations, we generate approaches not only likely to induce high benefits without increasing costs but also free of assumptions: we measure bio-geography as it is.

*Implications of all the available evidence:* The fact that asymptomatic cases were not tested in many countries may explain the exponential growth and much higher number of deaths observed in those countries. Ineffective testing (and, therefore, ineffective isolation) can also result from the absence of geo-referenced data analysis. Because the geographical location where people reside, work, study, or shop is not a random event, the analysis of small greographical areas is essential. Only when actual geographical relationships are observed, optimal (cost-benefit oriented) testing policies can be devised.

## Introduction

Albert Bartlett partially dedicated his life to teaching the expressions and consequences of the exponential function [1]. Epidemics –including the COVID-19 pandemic-tend to grow or decline exponentially. Given the catastrophic consequences of the epidemic exponential growth [2 Rudan], the first priority in epidemic control is to avoid contacts: when it takes just 60 days to increase from 1 to > 60,000 COVID-19 related deaths (as observed in this pandemic), ‘coexistence with the virus’ or ‘flattening the epidemic curve’ is not a likely outcome.

Because blocking contacts between infected and susceptible individuals prevents the exponential growth of an epidemic, testing is the first priority of a control policy: it allows to detect, treat if needed, and isolate all infected individuals. Hence, the WHO summarizes its recommendations with three words: *test, treat, isolate* [3].

However, a substantial proportion of individuals infected with SARS CoV-2 may be infectious even without symptoms. Hence, identifying asymptomatic cases is a crucial element in epidemic control: if they were rapidly identified and subsequently isolated, the virus could be removed from the environment and the pandemic would stop [4-7]. The theoretical solution for this problem is to test every inhabitant of every country.

Unfortunately, massive testing (the type required to identify all infected individuals) is not currently feasible in most countries. This situation creates a new challenge: how can major disseminators of the COVID-19 pandemic be identified when testing is limited (or very limited)?

Therefore, a fourth action should be added to the three proposed by WHO: *identification*. The determination of the geographical sites where asymptomatic cases are likely to be located is the linchpin of epidemic control. However, only a minor proportion of the population is tested on a given day (usually less than four per thousand inhabitants [8]). Given the scarce resources available, the limited testing, and the need to remove viral spreaders before the epidemic exponential growth overwhelms the anti-epidemic responses, identification should be precise, rapidly implemented, and cost-effective [9-11]. To address this composite problem, new metrics and methods may be required.

Test positivity (TP) is a concept to be considered. TP is the percentage of infections detected among tested individuals. While TP has been used for more than seven decades, in the context of COVID-19 it was first mentioned on March 30, 2020, by WHO [12]. Classically, TP has been utilized in reference to the ability of a test to identify and distinguish infection-positive from infection-negative individuals (also known as sensitivity and specificity,when the inference refers to the past; or test positive/negative predictive value, when the inference refers to the future). Such tests have a *diagnostic* purpose. They differ from *screening* tests [7, 13].

In this report, TP is viewed as a metric that may evaluate the efficacy of *testing programs* aimed at detecting asymptomatic COVID-19 cases; i.e., test positivity percentages may indirectly help to elucidate whether testing is or is not capturing asymptomatic cases, even when the number of tests performed per million inhabitants is low. In this context, TP is a component of an *epidemiologic* strategy used in a rapidly changing environment with the purpose of detecting and isolating infected individuals before the epidemic exponential process consolidates.

To the best of our knowledge, the validity of the TP metric to detect asymptomatic cases when testing is limited has not yet been demonstrated. To understand why TP percentages may achieve that goal, the history of epidemics is worth revisiting. When disease prevalence is zero (before an epidemic starts), the TP % is zero, too. The TP % is expected to be non-zero (but relatively low) after epidemic onset. That is so because even the worst epidemics on record have infected only a minor percentage of the population –for instance, the highest estimates on fatalities induced by the 1918 flu pandemic are about 5% [14].

Accordingly, when the TP % approaches or exceeds 50% (as reported in several countries affected by COVID-19), it is unlikely that such numbers reflect the actual disease prevalence. Instead, high TP percentages may express testing conducted within a subpopulation that does not (or only marginally) promote(s) epidemic spread: the group of *symptomatic* cases. Because symptomatic cases are easily detected, they are also easily treated and isolated. Consequently, symptomatic cases are not likely to disseminate epidemics. In contrast, infected but undetected individuals –asymptomatic cases-transmit the disease [15].

Therefore, *high TP* percentages may be observed when the testing policy predominantly detects *symptomatic* cases (as explicitly pursued by official policies or promoted by research institutions of some countries [16-18]). This hypothesis is further supported when the alternative is associated with promising outcomes, that is, when lower TP values are found in countries where the testing policy explicitly captures asymptomatic cases and, later, much fewer deaths per million inhabitants are reported than in countries reporting high TP percentages. This means that low TP percentages may reflect the prompt removal of SARS CoV-2-positive individuals –a likely outcome of policies that test asymptomatic cases. Thus, empirically elucidating whether high TP percentages are found when asymptomatic cases are excluded in the testing policy as well as the alternative hypothesis -low TP percentages are associated with testing programs that detect asymptomatic cases- are pre-requisites of scientifically sound testing policies meant to identify and isolate the spreaders of epidemics.

Other metrics of potential interest include: (i) the number of new cases, (ii) the number of COVID-19 related deaths/million inhabitants (deaths/mi), and (iii) the ratio between the number of tests and the number of cases detected (the T/C ratio). The time when the highest number of new cases occur is a relevant input of epidemiologic decision-making [19]. Because testing policies that only emphasize symptomatic cases will generate biased estimates on when new cases peak, approaches that detect asymptomatic cases are needed [16].

To demonstrate that novel testing methods can capture asymptomatic cases even when testing is limited, high-resolution, geo-referenced data may be required. When small geographical areas report high TP percentages, intensive testing in such areas could induce benefits that will include the larger surrounding area, generating high benefit-cost ratios [20].

Accordingly, relationships between the TP percentage, its inverse (the number of tests conducted per each detected case ratio or T/C ratio), and other metrics (the number of new cases and the number of deaths per million inhabitants) were investigated across countries and -using geo-referenced data of high resolution - across municipalities. This study aimed at elucidating whether, when and where the TP percentage and/or the T/C ratio informed earlier and/or provided more information than classic metrics in reference to asymptomatic cases and, if so demonstrated, whether such metrics, once geo-referenced, could foster region-specific, cost-benefit oriented control policies.

## Materials and methods

### Methods

Data on new (daily) tests performed, cases identified, test positivity (the percentage of positive cases/tests performed) and its inverse (the test/case ratio), as well as the cumulative number of tests or deaths per million inhabitants were collected from publicily available data sources and compared across countries and across time. Countries were classified as either displaying high or low test positivity (TP) when, at least three months after the initiation of the epidemic, they showed non-overlapping intervals of TP and such classes also revealed non-overlapping intervals of epidemic-related outcomes (deaths/million inhabitants). Differences in median deaths/million inhabitants between low TP and high TP countries were investigated with the Mann-Whitney test using a commercial statistical package (Minitab LLC, State College, PA, USA). Geographical data were processed with ARC GIS software (ESRI, Redlands, Ca, USA). Thirteen countries were evaluated because, together, they exhibited geographical and demographic diversity and individually, they were affected by COVID-19 long enough (two or more months) so they could reveal exponential epidemic growth, if it happened. Such countries were: Bangladesh, Belgium, Colombia, Cyprus, Ethiopia, Germany, Greece, Italy, South Africa, South Korea, Spain, Uruguay, and the United States. To explore georeferenced data on test positivity, the municipality-specific data reported by Harvard University on Puerto Rico were analyzed [https://rconnect.dfci.harvard.edu/covidpr/?s=08]. Because many countries have reported errors and/or changed the definition of cases and deaths and because not all affected countries were investigated, inferences should not be construed to be representative. However, inferences derived from patterns observed in two or more countries (or regions of a country) may be viewed as plausible and, possibly, generalizable.

### Data

Data were extracted from *Worldometer* and/or governmental sites (

https://www.worldometers.info/coronavirus/; https://www.pio.gov.cy/coronavirus/eng;

https://eody.gov.gr/epidimiologika-statistika-dedomena/ektheseis-covid-19/;

https://www.gub.uy/sistema-nacional-emergencias/).

## Results

### Test positivity-related temporal patterns

Findings revealed two temporal patterns: (i) one characterized by an early TP peak (the highest TP percentages took place in the first weeks), followed by a rapid decline to ∼ 1% (four countries, green rectangles, Figs. 1 A-D); and (ii) a pattern shared by six countries, which showed later and increasing TP values, even >10%, over one or more weeks (red rectangles, Figs. 1 E-J). In addition, the data of ten countries consistently demonstrated that the highest TP percentages occurred earlier or at the same time as, but not later than the number of new cases (Fig. 1 A-J). The first pattern (low TP percentage) was associated with a quasi-complete cessation of the epidemic within four months. The second pattern (high TP percentages) was shown by countries where, four to six months into the epidemic, the median number of deaths/mi was at least 10 times higher than in countries displaying the alternative pattern (*p*<0.02, Fig. 2).

**Fig. 1.**
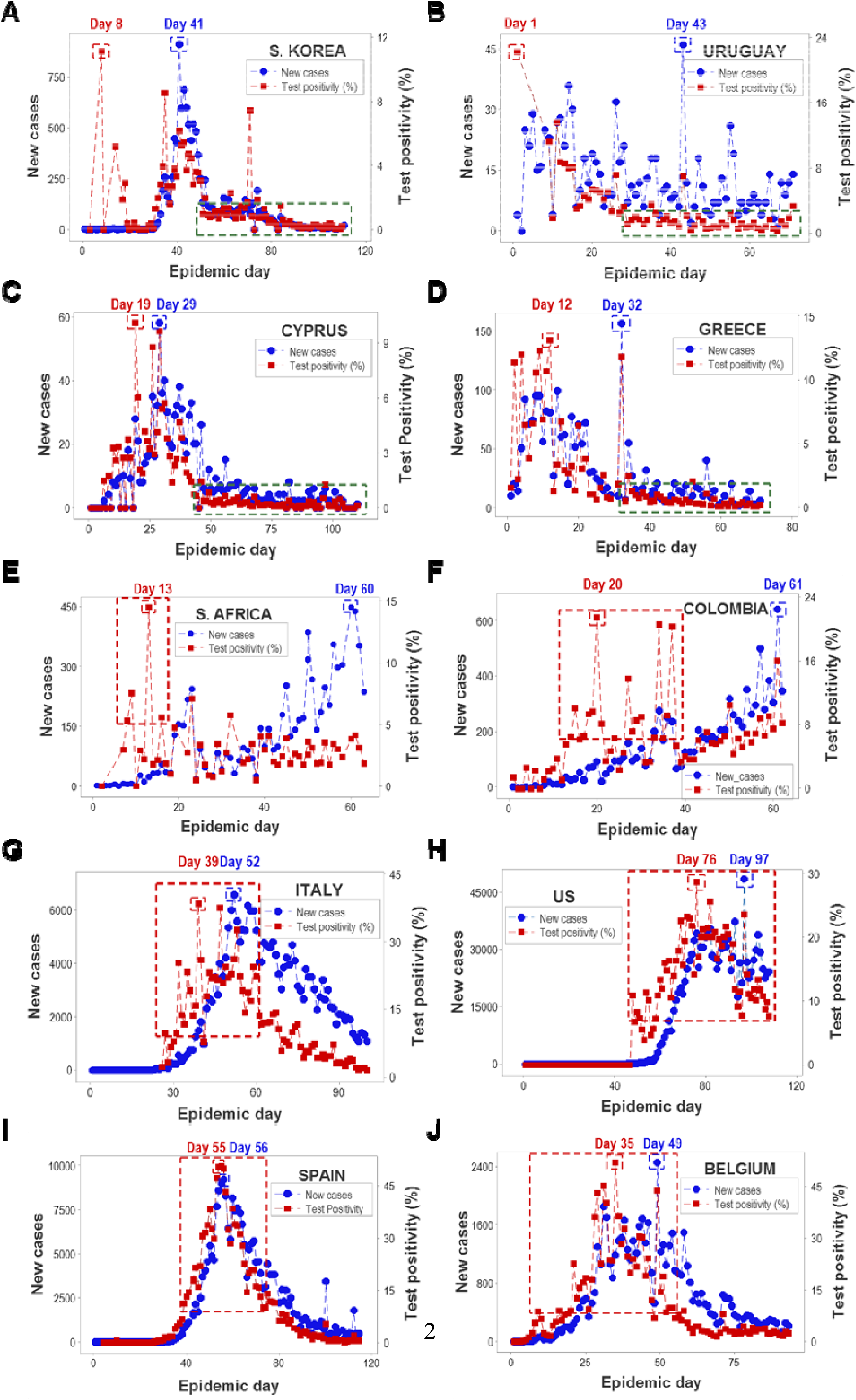
Temporal relationships between the test positivity percentage and the number of new cases. Two epidemic patterns were differentiated among10 countries: (i) epidemics that revealed a rapid decrease in the test positivity (TP) percentage and, subsequently, displayed very low (<1%) TP values (green rectangles, **A-D**); and (ii) epidemics that reported either later TP peaks, increasing percentages of the TP over two or more weeks, and/or TP percentages higher than 5% (red rectangles, **E-J**). In all ten countries investigated, the peak TP percentage occurred earlier than the peak number of new cases. Source: *Worldometer*.

**Fig. 2.**
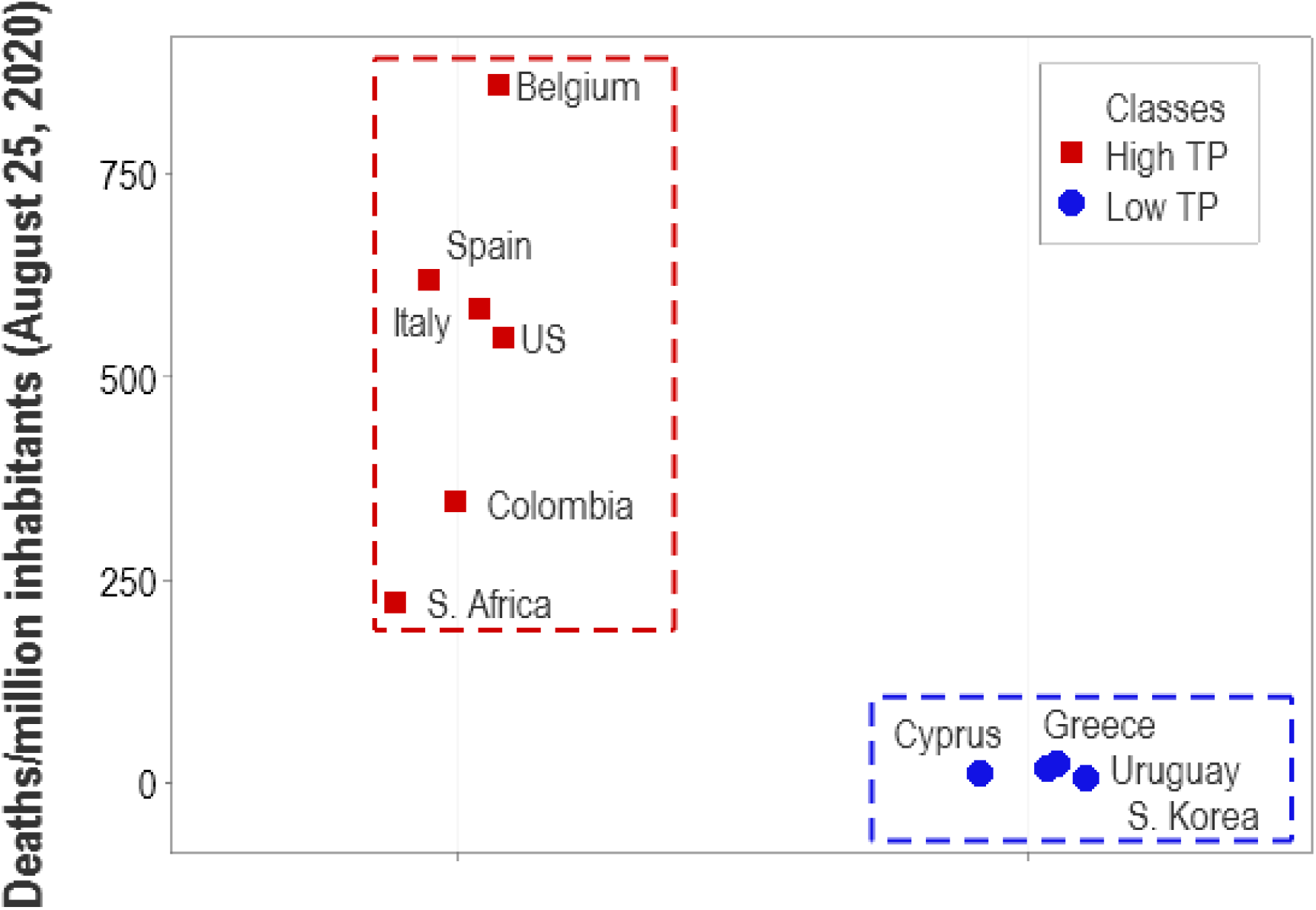
Temporal relationships between test positivity percentage and deaths per million inhabitants. By August 25, 2020 (at least four months into the epidemic), the group of countries that, earlier, reported high (three-digit) test positivity percentages (high TP) showed at least 10 times more deaths per million inhabitants (d/mi) than countries that, earlier displayed low (two-digit) TP percentages (*p*< 0.02, Mann-Whitney test). High and low TP countries revealed non-overlapping data intervals (rectangles). Source: *Worldometer*.

### Validation of TP-based testing policies designed to detect asymptomatic cases

Four-dimensional (3D and temporal) plots that included TP percentages and T/C ratios identified three profiles, characterized by: (i) high TP percentages (data points shown only in the upper right quadrant, 4 countries); (ii) a hybrid pattern, which exhibited high TP and some high T/C observations (one country); and (iii) countries predominantly displaying high T/C ratio values (four countries, Figs. 3 A-I). TP values less than 5 % did not predict the end of the epidemic –even countries displaying < 1% TP over more than two weeks failed to report zero cases after such a period of time (Figs. 4 A-D).

**Fig. 3.**
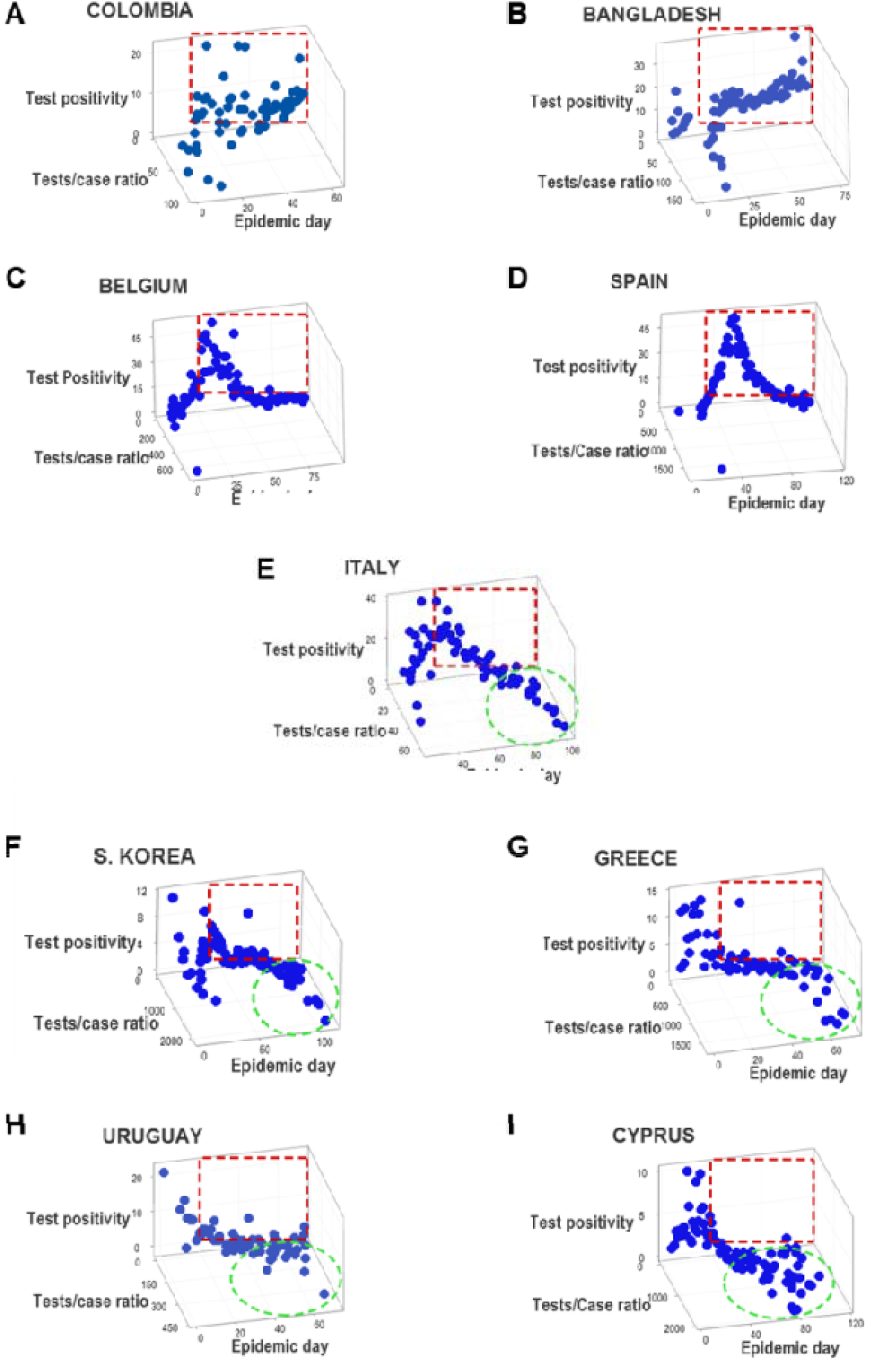
Temporal relationships between test positivity and the tests/case ratio. The assessment of the tests/case ratio revealed more information when nine countries were investigated, which partially included the set reported in Fig. 1. Four countries showed high TP percentages but few or no high tests/case ratios (**A-D**). One country displayed a hybrid pattern, which was characterized by a few high TP percentages and a few high tests/case ratios (**E**). The remaining four countries predominantly exhibited high tests/case ratios (**F-I**).

**Fig. 4.**
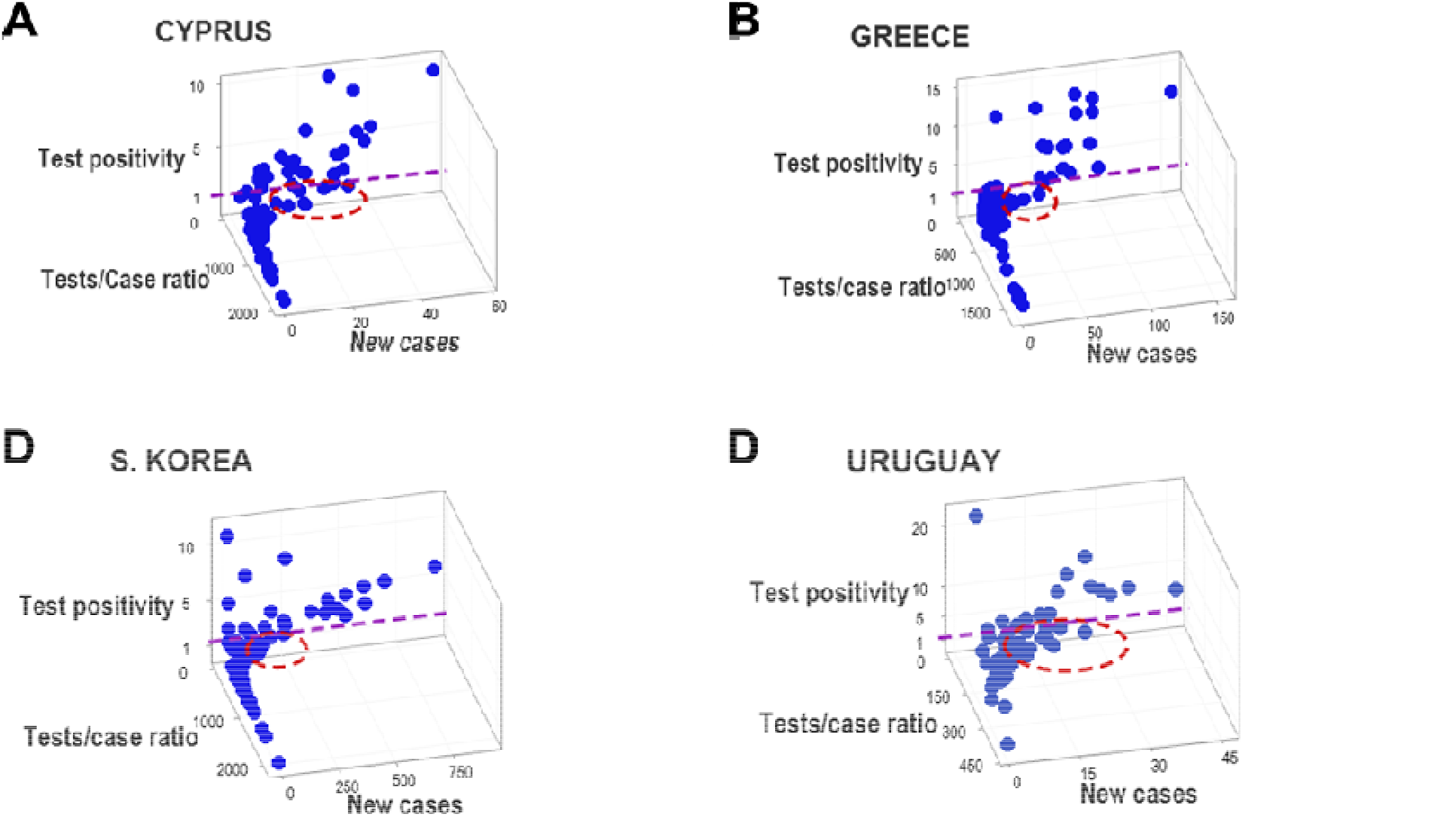
Pattern- vs. threshold-based evaluations of epidemic control measures. Four countries located in three continents showed new cases even when the percentage of test positivity (TP) was < 1% (ovals, **A-D**). Because new cases were observed over protracted (more than two-week long) periods of time, it is concluded that no numerical cutoff (e.g., TP < 5%) indicates an epidemic is under control. In contrast, patterns –such as a perpendicular data inflection– promote inferences, regardless of any cutoff value.

### Exploration of geo-referenced and temporal TP as tools of cost-effective policies

To investigate the hypothesis that geo-referenced and temporal TP data support site-specific, cost-effective interventions, the test positivity percentages reported in all municipalities of Puerto Rico on September 11, 2020, were investigated. One municipality reported a TP >50%, which was surrounded by 17 municipalities that reported much lower TPs. Assuming that testing resources (including tests) were distributed according to the size of the population, the municipality with >50% TP received 1/42 of such resources and the remaining municipalities obtained testing-related resources 42 times larger in magnitude (Fig. 5).

**Fig. 5.**
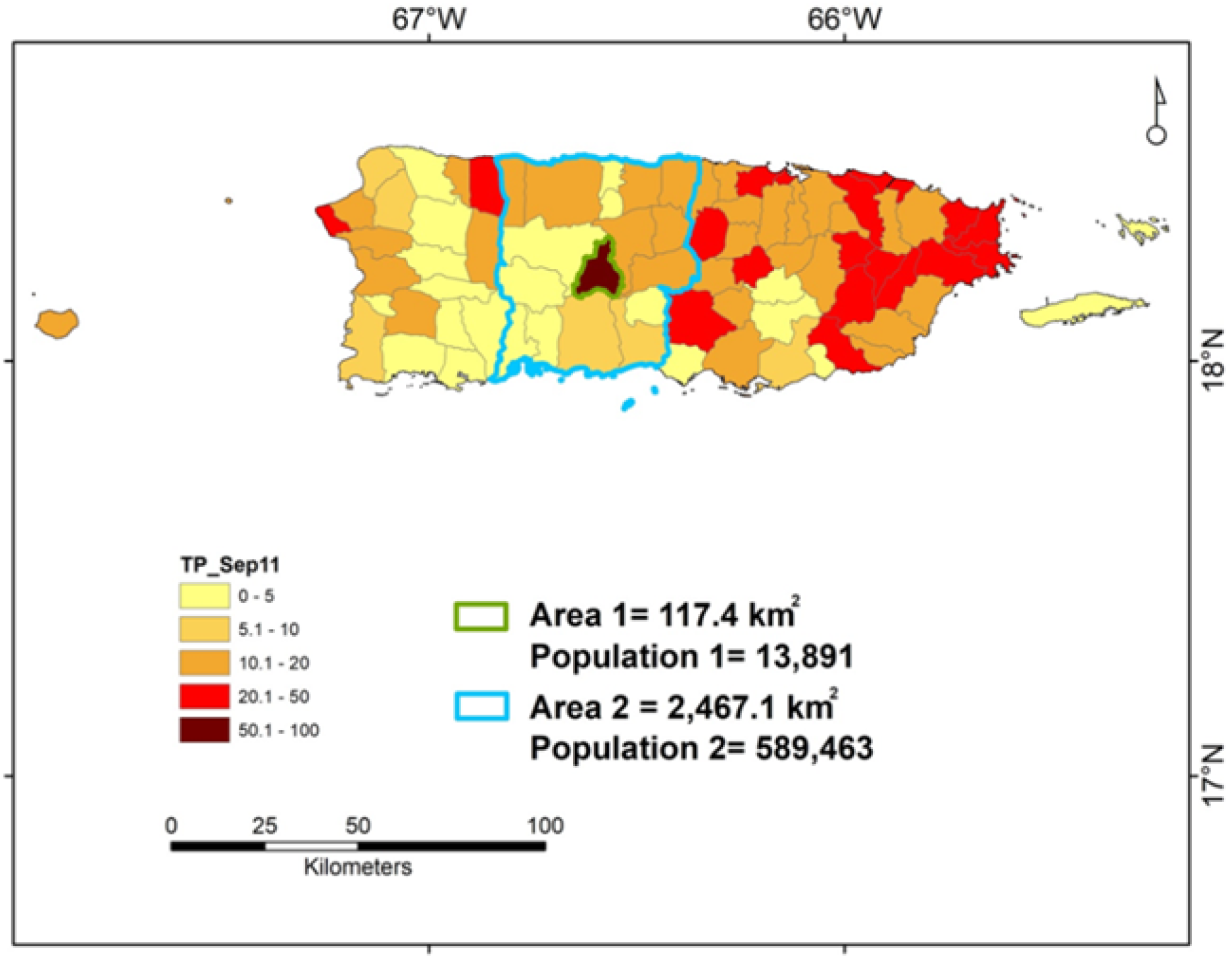
Geo-epidemiologically specific, cost-benefit effective, differential testing. The test positivity (TP) percentages reported in all municipalities of Puerto Rico, on Sept 11, 2020, are depicted. Area 1 shows the municipality of Jayuya. Area 2 identifies a larger, surrounding region that includes ∼42 times more people and covers an area ∼ 8.5 times larger than those of Jayuya. Given Jayuya +50% TP, a greater testing effort conducted in this municipality may yield large benefits, which may also include the surrounding area (Area 2) and, indirectly, benefit the western half of the island. Vice versa, keeping the same level of testing performed before may lead to long-lasting, costly consequences: if the virus circulating within Jayuya reached the surrounding area and the prevalence of COVID-19 became similar to the one affecting Jayuya (which is estimated by test positivity), then a much larger number of people (up to 42 times larger) could become infected, who would reside in an area 8.5 times larger, i.e., control would then be much harder, longer and costlier.

Such a situation facilitated cost-benefit oriented, geographically and epidemiologically specific interventions. For instance, if the level of testing implemented in area 1 (the municipality with >50% TP) was decided to be increased 1000%, immediately -but no new resources were provided-, the additional tests required in area 1 could only come from area 2. This means that 10 of the original 43 units of resources spent in areas 1 and 2 (1 unit spent in area 1 and 42 units spent in area 2) would now be used in area 1 (a 1000% increase). This solution would cause a 23.56 % reduction (from 42 to 32) in the units of resources spent in area 2 - a region barely affected by the epidemic and, therefore, highly vulnerable (Fig. 5).

These data also estimated the costs of random testing and/or testing that ignores geo-temporal cost-benefit considerations. If the surrounding area (area 2) was affected as much as area 1, the size of epidemic could increase 42 times (Fig. 5). This epidemic growth could occur even if testing continued, uninterruptedly, at the same level as previously performed.

### Additional tools

Relationships between metrics elucidated when testing was or was not adequate. Examples collected from 14 countries revealed when testing grew faster than cases (a desirable outcome) or the alternative pattern suggested additional testing was needed (Fig. 6A-D). The use of the number of deaths per million inhabitants was also informative (Figs. 6 E-H).

**Fig. 6.**
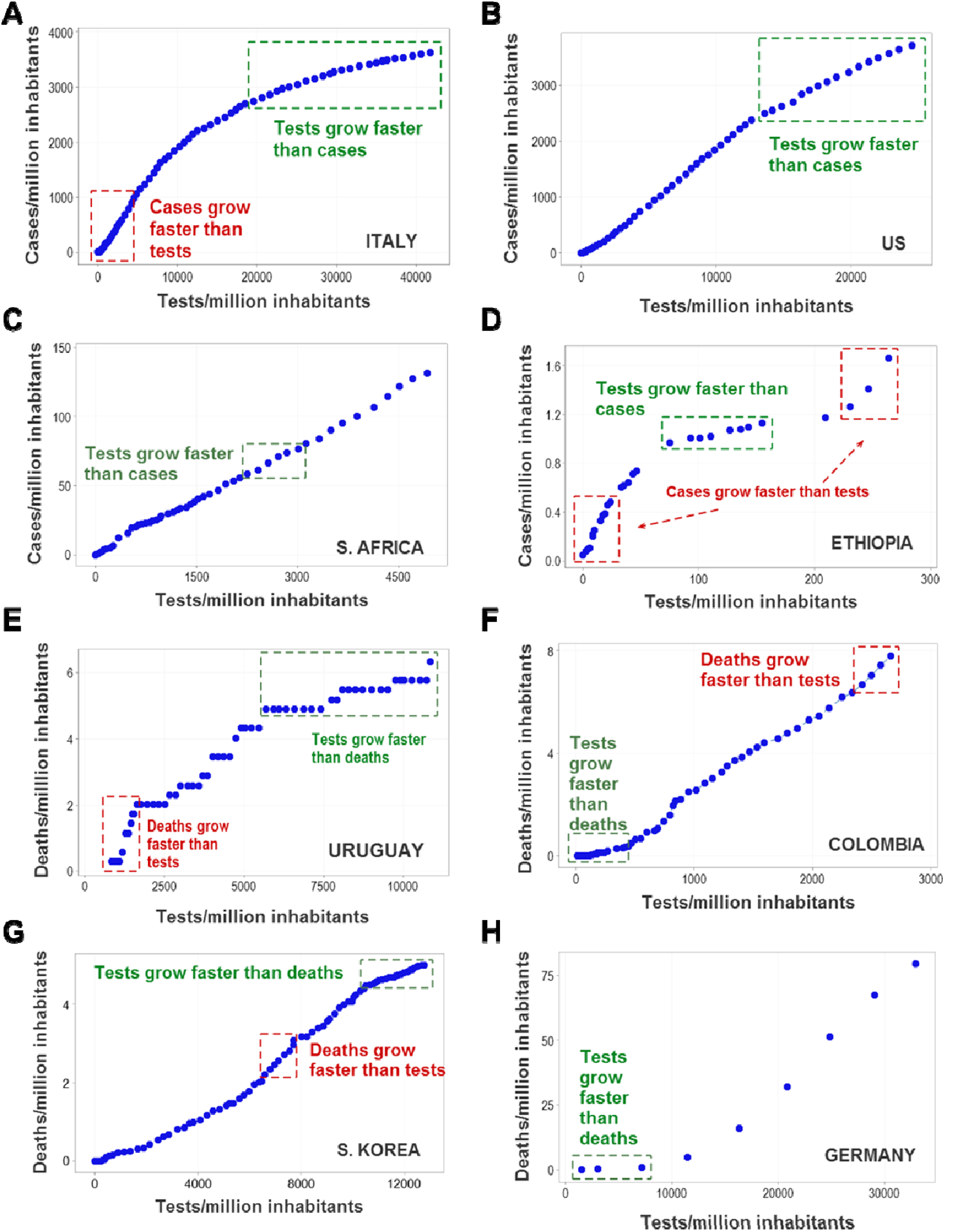
Relationships between outcomes (non-fatal or fatal infections) and the number of tests/million inhabitants. Testing policies may be adjusted based on patterns generated by cases (**A-D**) and fatalities (**E-H**). When testing grows slower than cases or deaths (when the curvature approaches a vertical pattern), testing is likely to be insufficient or inadequate.

## Discussion

Because asymptomatic patients rarely request medical assistance, they may become the invisible spreaders of epidemics. Consequently, testing, treating, and isolating such cases should be prioritized [3]. However, given that testing is usually limited, their identification is problematic [8]. This compounded problem was addressed with an approach that, considering test positivity (TP), evaluated testing policies that prioritize the detection of asymptomatic cases and also considers geo-referenced, cost-benefit oriented data analysis.

Testing symptomatic or asymptomatic cases may depend on medical and public health-related perspectives [7]. For instance, testing that emphasizes symptomatic cases differs from screening-oriented testing. In the first case, it is assumed that epidemics can be stopped through interventions conducted within hospitals –as when individuals that feel ill seek medical assistance. It is expected to find high percentages of test positivity when *confirmatory* testing is promoted (the type of testing physicians are likely to prescribe). While this type of testing (or purpose) is necessary when a clinical perspective is considered, it does not contribute much to prevent epidemic spread, which depends on early detection of asymptomatic cases [7,18]. In contrast, in population medicine, *epidemiologic* testing is needed (screening or information usable in epidemic control, not only in clinical medicine). Here it was hypothesized that lower TP percentages predicted lower numbers of deaths per million inhabitants than when higher TP percentages were observed –findings consistent with the hypothesis that asymptomatic cases are detected when the percentage of TP is low. Findings supported these hypotheses, opening a path toward scientifically grounded testing policies that seek to detect (and immediately isolate) asymptomatic –not only symptomatic-cases [7, 18]. As expected, countries that displayed high TP percentages in early epidemic stages, later reported many more deaths/mi than low TP countries. These findings supported the notion that, in high TP countries, asymptomatic cases were missed and, consequently, the epidemic grew exponentially and so did deaths.

The epidemics here investigated also showed that TP peaks occur earlier than the peak number of new cases. Data turning points (which include peaks) help to conduct epidemiologic forecasts [21].

Findings also addressed the statement made by WHO on May 12, 2020, which suggested that low (<5%) TP values over two consecutive weeks signal that an epidemic is under control [22]. This study found that even 1% TP was associated with longer epidemics (Figs. 3 A-D).

The fact that neither a 5% nor a 1 % TP cutoff predicted the immediate cessation of an epidemic was expected: dichotomization of continuous data promotes errors. Such a procedure assumes that the discrete (discontinuous) nature of a process can change below or above a cutoff generated by continuous data. Yet, the evidence demonstrated that an ‘epidemic-positive’ country does not become an ‘epidemic-negative’ one when TP changes from 5.00 to 4.99 %. While pattern recognition can discriminates without numerical cutoffs (as Fig. 5 shows), the dichotomization assumption tends to lose information and generates errors [23].

Therefore, TP seems a robust and explanatory metric. Unlike the number of new cases, TP may inform *when* (and, provided that geo-referenced information is also available, *where*) spikes of its percentage will identify specific places that require additional testing so asymptomatic cases can be rapidly detected, isolated, and treated [24]. However, to prevent such patients from promoting exponential epidemic growth, early and geographically-specific responses are needed. Here, an operational model showed that observable geo-epidemiologic features can generate site-specific, cost-effective interventions.

Investigating geo-referenced epidemiologic data matters because humans are not homogeneously distributed in space. Lakes, railroads, schools and many other (geographically specific) entities both separate and connect humans. While some geographical points act as epidemic nodes, many points do not seem to influence epidemic spread [25].

While earlier studies have conducted cost-benefit analysis of COVID-19 control policies, they lacked geo-referenced data [26]. The geo-referenced, cost-benefit theory followed in this study assumed that epidemics disseminate as floods do: from a ‘higher’ point (where disease prevalence or its surrogate, test positivity, is higher) toward surrounding areas where test positivity percentages are lower. When cost-benefit oriented interventions are not immediately implemented in such situations, the epidemiologic condition of the surrounding region can rapidly worsen. This epidemiologic ‘flood’ metaphor was empirically supported: rapid interventions in small areas that threaten larger areas tend to be highly cost-effective and may stop epidemics (Fig. 4).

While aggregate (state- or country-level) data do not reveal spatial relationships and, therefore, are not very useful [27], non-aggregate geo-referenced information –e.g., data on neighborhoods- are epidemiologically usable, as shown with municipality-level information collected in Puerto Rico (Fig. 5). When geographical-epidemiological relationships are considered, it is possible to identify policies that benefit not only the area of direct intervention but also the surrounding area –as commonly practiced in flood protection-related policies [28]. This approach has also been described as the ‘wrong pocket problem’, i.e., the area where the investment is made does not only benefit from it: a larger area is benefited as well [29]. One example of this geographically grounded, cost-benefit oriented approach has recently been applied in an emergency vaccination against rabies conducted in Tanzania, where a limited intervention in a small ring that surrounded a large city also benefitted the large city [11].

Findings support the view that, to stop epidemic exponential growth, testing policies should aim at detecting all infected individuals (including asymptomatic cases). In spite of its relevance, testing policy seems an area that lacks interdisciplinary scientific expertise. Approaches that include temporal and geo-referenced test positivity data, as well as cost-benefit oriented considerations, may contribute toward filling that gap.

## Data Availability

All data utilized are publicly available.

## Contributors

All authors contributed to model design, interpretation of results, and writing of the report. ALR, ALH, AI, AA and PV collected data. JBH and FOF transcribed and analyzed the data. JLF made maps, ALR and MHVvR drafted the manuscript.

## Data sharing

No primary data were collected for this study, which used publicly available data. The sources of the data used are described in this article.

